# An assessment of a rapid SARS-CoV-2 antigen test in Bangladesh

**DOI:** 10.1101/2021.10.05.21264551

**Authors:** Zannat Kawser, Mohabbat Hossain, Sara Suliman, Shahin Lockman, Jesse Gitaka, Gama Bandawe, Redwan Rahmat, Imrul Hasan, Abu Bakar Siddik, Mokibul Hassan Afrad, Mohammed Ziaur Rahman, Glenn Miller, David R. Walt, Louise C. Ivers, Regina C. LaRocque, Jason B. Harris, Firdausi Qadri

## Abstract

Early detection of SARS-CoV-2 infection is crucial to prevent the spread of the virus. In this study, we evaluated the performance of a commercial rapid antigen detection test, BD Veritor, and compared this (and another rapid test, Standard Q) against a gold-standard of nasopharyngeal (NP) swab tested by reverse transcription-polymerase chain reaction (RT-PCR) in prospectively-recruited adults in Dhaka, Bangladesh. We compared the sensitivity and specificity of the two rapid antigen tests against RT-PCR results in 130 symptomatic and 130 asymptomatic adults. In addition, we evaluated the suitability and ease-of-use of the BD Veritor test in a subsample of study participants (n=42) and implementers (n=5). The sensitivity of the BD Veritor rapid antigen test was 70% in symptomatic (95% confidence interval [CI]: 51-85%) and 87% (95% CI: 69-96%) in asymptomatic individuals with positive SARSCoV-2 RT-PCR, for overall sensitivity of 78% (95% CI: 66-88%). The sensitivity of the Standard Q rapid antigen test was 63% (95% CI: 44-80%) in symptomatic and 73% (95% CI: 54-87%) in asymptomatic individuals. One false positive in BD Veritor test (specificity 99.5) and no false positive in Standard Q tests were observed (specificity 100%). The BD Veritor rapid antigen test was 78% sensitive when compared with RT-PCR irrespective of the cycle threshold (Ct) levels in this evaluation in Bangladesh. The implementation evaluation data showed good acceptability in the field settings. This warrants large field evaluation as well as use of the rapid antigen test for quick assessment of SARS-CoV-2 for containment of epidemics in the country.

## Introduction

Rapid antigen detection tests are point-of-care immunochromatographic assays which detect protein antigens specific to the Severe Acute Respiratory Syndrome of the Coronavirus-2 (SARS-CoV-2) (e.g. nucleocapsid) (1). The ease-of-use and quick turnaround time of such tests can expand access to testing and decrease delays in diagnosis (2). Furthermore, modeling studies on SARS-CoV-2 have demonstrated that even if rapid antigen testing is associated with decreased sensitivity, the accessibility and short turnaround time in reporting results may be advantageous for decreasing transmission (3). Rapid antigen testing is particularly useful if deployed in the context of repeated testing over time (4) (5).

The performance of the rapid antigen tests has been determined by comparing their sensitivity and specificity with nucleic acid detection-based reference reaction (6). The current gold standard for identifying the presence of SARS-CoV-2 is reverse transcription-polymerase chain reaction (RT-PCR) in samples collected by nasopharyngeal (NP) swab (7). Despite their high sensitivity, nucleic acid amplification tests are associated with the need for laboratory processing, high costs, and a longer turnaround from sampling to return of results (8) (9). The NP swabs are also more challenging and uncomfortable (for patients) to collect than anterior nares swabs. For this reason, rapid antigen testing is a valuable tool for contact tracing and early detection of COVID-19 patients to triage for treatment options, especially in settings where RT-PCR is less available or where follow-up reporting of RT-PCR results is more difficult, and particularly when anterior nares samples can be used.

In this study among asymptomatic and symptomatic adults, we evaluated the performance (sensitivity/specificity) of two rapid antigen detection tests, the BD Veritor™ (Becton-Dickenson, USA) and the Standard Q™ (SD-Biosensor, Korea) rapid antigen test, in comparison to NP swab RT-PCR as the reference standard. The BD Veritor was performed according to the manufacturer’s recommendations using an anterior nares swab specimen, while the Standard Q and reference RT-PCR were performed on nasopharyngeal swab specimens. We also evaluated the performance of the rapid antigen tests across the spectrum of RT-PCR cycle threshold (Ct) values. Finally, we assessed the implementation characteristics of the BD Veritor rapid antigen test, including fitness-for-use in different populations and settings in Bangladesh.

## Methodology

### Study design and participants

We enrolled study participants at a triage and sample collection booth at Kurmitola General Hospital (n=49) as well as at the ideSHi COVID-19 testing facility (n=211) in Dhaka, Bangladesh. Adults aged 18 and above were eligible for inclusion. For this analysis, we aimed to enroll 130 symptomatic patients with Covid-19 like symptoms including fever, cough, headache, sore throat, shortness of breath and fatigue (10) who had their onset of first symptom within five days, including 100 individuals with negative RT-PCR results and 30 individuals with positive RT-PCR results. In addition, we aimed for a similar target for positive and negative asymptomatic individuals who presented for routine COVID-19 screening at the above sites (primarily occupational screening or for known contact with an individual who tested positive). Written informed consent was obtained from participants. The study was approved by the Research Review Committee (RRC) and Ethical Review Committee (ERC) of icddr,b (Protocol no: PR-20042).

### Specimen collection

Nasopharyngeal swab specimens were collected by trained personnel and placed in a 3-mL tube of viral transport medium (Citoswab, Citotest Labware Manufacturing Co. Ltd, China) to be used for both Standard Q antigen testing and RT-PCR testing. Anterior nares swab samples were also collected by trained personnel according to the manufacturers’ instructions for BD Veritor. Specifically, the swab provided with the kit was inserted into the anterior nasal cavity up to 2.5 cm and rolled 5 times along the mucosal surface in both nostrils. The nasopharyngeal and anterior nares swab specimens were collected simultaneously until the first 200 individuals with negative RT-PCR results were enrolled in each group (100 symptomatic and 100 asymptomatic). Thereafter, the anterior nares samples were collected within 24 hours of the nasopharyngeal specimen until 60 individuals with positive RT-PCR results were accrued (30 symptomatic and 30 asymptomatic).

### RT-PCR on nasopharyngeal swab specimens

Viral RNA was extracted from 200 ul of viral transport media using the magnetic bead based Nexor 32 Fully Automated Nucleic Acid Extractor (Nucleic Acid Extraction or Purification Kit, Beijing Lepu Medical Technology Co., Ltd, China). RT-PCR was carried out using the China CDC primer and probes. In brief, this was performed in a 20 μl reaction volume and each reaction contained extracted RNA, 2x iTaq Universal Probes Reaction Master Mix (Biorad, CA, USA), iScript Reverse Transcriptase, the CDC_ORF1ab and N forward and reverse primers, and probe (11)(12). Specimens were determined to be positive for SARS-CoV-2 when the ORF1ab and N genes were detected with an exponential growth curve and a cycle threshold (Ct) value <40, and negative when these genes could not be detected. The test was considered positive even if one gene was detected. The quality of the nasopharyngeal specimen extracted was determined by analyzing the curve generated with the *rnaseP* housekeeping gene.

### Rapid Antigen Testing

The rapid tests were performed in accordance with the manufacturer’s instructions. For the BD Veritor assay, the anterior nares swab was inserted into the extraction reagent tube and mixed in the fluid for a minimum of 15 seconds before discarding. Three drops of the processed specimen were added to the sample well of the device, and incubated for 15 minutes. Following this, the test device was inserted into the Veritor Plus Analyzer (BD) for reading.

For the Standard Q kit, 350 μL of freshly obtained NP swab specimen in viral transport medium was reconstituted in the extraction buffer supplied by the manufacturer and incubated for 45-50 minutes. For testing, 3 drops (approximately 80 μL) of extracted nasopharyngeal specimen was applied to the sample well of device, and results were interpreted after 15 minutes, based on the manufacturer’s instructions.

### Assessment of implementation characteristics

We surveyed 5 test implementers and 42 participants about the BD Veritor test with a user acceptability and adoption assessment form for implementers and a feedback form for participants. Five point Likert scale was used for documenting the level of satisfaction and level of difficulties in addition to the qualitative aspects in the questionnaire. We also assessed the BD Veritor test in comparison to NP swab RT-PCR regarding resources to collect and transport samples, and use of Personal Protective Equipment (PPE) and consumables. We evaluated turnaround times (from the sample collection time until the result was reported to the participant) for each sample type, assay, and platform from the time of collection to delivery of results.

### Statistical analysis

We calculated the sensitivity and specificity (13) of each rapid test compared with the NP RT-PCR gold standard, and reported these as a percentage with 95% confidence intervals (CIs). The sensitivity of both rapid tests (combined) was also analyzed in RT-PCR positive samples stratified by Ct value >30, 20-30, and <20. The sensitivity of the two rapid tests was compared and P value was calculated using McNemar’s Chi-Square Test. In addition, the comparison of Ct values between symptomatic and asymptomatic individuals was performed using the Mann-Whitney U test.

## Results

We enrolled 262 individuals in this study. Two study participants were subsequently excluded from the analysis because of incomplete information in the case record forms, resulting in a study set of 260 individuals. Forty-nine of the symptomatic individuals were enrolled at the Kurmitola General Hospital, while the remaining 211 participants included in the analysis were enrolled at the ideSHi testing facility. Demographic characteristics are shown in Table 1. The median age of the symptomatic individuals was 35 years (range, 18 – 81 years), and 53% were male. The most common symptom was fever (N=88, 68%), and the median duration (and range) of symptoms were 3 days (1 to 5 days) in the symptomatic patients. The median age of the asymptomatic individuals was 33 years (range, 18 – 74), and 81% were male.

**Table 1:** Demographic characteristics in different groups of participants.

| Characteristic | Symptomatic |  | Asymptomatic |  |
| --- | --- | --- | --- | --- |
|  | PCR+(n=30) | PCR-(n=100) | PCR+(n=30) | PCR-(n=100) |
| Median Age | 46.5 | 32 | 33.5 | 33 |
| **Sex** |  |  |  |  |
| Male | 15(50%) | 54(54%) | 29(96%) | 76(76%) |
| Female | 15(50%) | 46(46%) | 1(4%) | 24(24%) |
| **Population category** |  |  |  |  |
| Traveler | 0 (0%) | 0 (0%) | 19 (63.3%) | 27 (27%) |
| Student | 4 (13.3%) | 19 (19%) | 2 (6.7%) | 1 (1%) |
| Healthcare worker | 3 (10%) | 6 (6%) | 0 (0%) | 2 (2%) |
| General population | 23 (76.6%) | 75 (75%) | 9 (30%) | 70 (70%) |
| **\*\*Symptom spectrum** |  |  | - | - |
| Fever alone | 1(3.3%) | 13 (13%) |  |  |
| Fever+Cough | 4 (13.3%) | 7 (7%) |  |  |
| Fever+Headache | 0 (0%) | 5 (5%) |  |  |
| Fever+Cough+Headache | 5 (16.7%) | 10 (10%) |  |  |
| Symptoms without fever | 7 (23.3%) | 35 (35%) |  |  |
| **History of contact** | 15 (50%) | 43 (43%) | 7 (23.3%) | 3 (3%) |
| **History of regular use of mask** | 27 (90%) | 89 (89%) | 24 (80%) | 97 (97%) |

\*Health care worker; \*\*Other symptoms not mentioned indicate sore throat, shortness of breath, diarrhea, loss of taste and/or loss of smell, generalized weakness

The sensitivity and specificity of both rapid tests are reported in Table 2. The sensitivity of the BD Veritor test (78%) was higher than that of the Standard Q test (68%) (P=0.041). The sensitivity of both tests was higher in asymptomatic individuals than in symptomatic individuals. Only one false positive BD Veritor test was identified in a screened symptomatic individual who reported a four-day history of cough with no fever, myalgia or loss of smell. No false positive Standard Q tests were observed.

**Table 2:** Performance of two rapid antigen detection tests for detection of SARS-CoV-2 in comparison to nasopharyngeal RT-PCR.

| Category |  | *BD + (n) | BD - (n) | BD performance (95% CI) | *Standard Q + (n) | Standard Q - (n) | Standard Q performance (95% CI) |
| --- | --- | --- | --- | --- | --- | --- | --- |
| Symptomatic | PCR + (N=30) | 21 | 9 | Sensitivity 70% (54-88%) | 19 | 11 | Sensitivity 63% (47-83%) |
|  | PCR - (N=100) | 1 | 99 | Specificity 99% (95-100%) | 0 | 100 | Specificity 100% (69-100%) |
| Asymptomatic | PCR + (N=30) | 26 | 4 | Sensitivity 87% (69-96%) | 22 | 8 | Sensitivity 73% (61-92%) |
|  | PCR - (N=100) | 0 | 100 | Specificity 100% (69-100%) | 0 | 100 | Specificity 100% (69-100%) |
| **Overall sensitivity and specificity (both symptomatic and asymptomatic participants)** |  |  |  |  |  |  |  |
| **Positive PCR** | N = 60 | 47 | 13 | Sensitivity 78% (66-88%) | 41 | 19 | Sensitivity 68% (55-80%) |
| **Negative PCR** | N= (200) | 1 | 199 | Specificity 99.5% (97-100%) | 0 | 200 | Specificity 100% (98-100%) |
| **Combined symptomatic and asymptomatic categorized by RT-PCR cycle threshold (Ct) value** |  |  |  |  |  |  |  |
| Ct > 30 | N = 13 | 1 | 12 | Sensitivity<br>8%<br>(0-36%) | 0 | 13 | Sensitivity<br>0%<br>(0-25%) |
| Ct 20-30 | N = 32 | 31 | 1 | Sensitivity<br>97%<br>(84-100%) | 26 | 6 | Sensitivity<br>81%<br>(64-93%) |
| Ct < 20 | N = 15 | 15 | 0 | Sensitivity<br>100%<br>(78-100%) | 15 | 0 | Sensitivity<br>100%<br>(78-100%) |

\*For BD Veritor™ testing kit, anterior nares specimens and for Standard Q, nasopharyngeal
specimen were tested based on manufacturer's instruction.

We compared the performance of both rapid antigen tests stratified by the observed Ct value of the nasopharyngeal RT-PCR. There was no difference in the median Ct value between the symptomatic and asymptomatic individuals (Figure 1). We also observed no significant difference (P=0.2 by Mann-Whitney U) in the range of Ct values between the two groups and hence considered such individuals together in the analysis stratified by Ct value (Table 2). For RT-PCR samples with a Ct < 20 (i.e., those with a high amount of viral genetic material), the sensitivity of both rapid antigen tests was 100%. For RT-PCR samples with a Ct > 30 (i.e., those with little viral genetic material), neither rapid antigen test performed well (sensitivity <10%). For RT-PCR samples with Ct values in the 20-30 range, the sensitivity of the BD Veritor test was 97% (95% CI: 84-100%), compared with 81% (95% CI: 64-93%) for the Standard Q.

**Figure 1.**
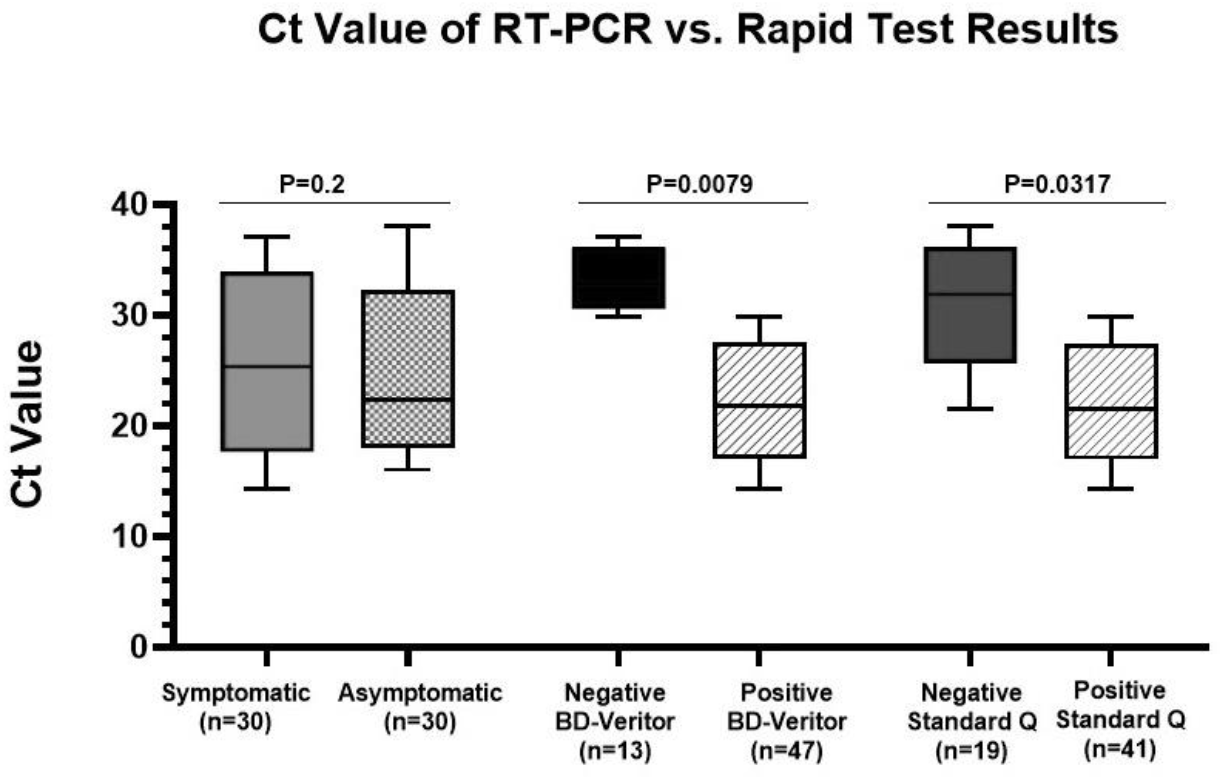
Comparison of Ct values among symptomatic and asymptomatic study participants, and across rapid antigen tests. The was no significant difference in the median Ct value in PCR positive symptomatic and asymptomatic participants, however, the median Ct values were significantly higher in individuals with false negative rapid antigen tests.

We also interviewed a subgroup of participants (n=42) within 24 hours after having obtained the anterior nares swab for the BD Veritor™ rapid test and asked them about the suitability of the test, the ease of the sample collection process, the ease of testing, the accuracy of the test and the turnaround time. Four participants were dissatisfied with the accuracy of the result by the BD Veritor™, while the remainder of the participants (n=38) were satisfied. All of the 42 interviewees expressed satisfaction with the sample collection process, ease of testing and the 15-20 minutes turnaround time. The 5 implementers surveyed, each of whom performed more than 50 tests, were satisfied with the kit components, the design of the device, the kit storage conditions, the quality controls, the time taken for the sequence of steps, the read-out of the results and the suitability for batch testing along with sequential testing. Based on their experience, testers estimated more than 100 patients could be tested and results given with the BD Veritor™ kit in one 8 hour period of a working day.

## Discussion

We evaluated the performance ofthe BD Veritor™ rapid antigen test and also compared with the Standard Q™ antigen test, for detecting SARS-CoV-2 in asymptomatic and symptomatic adults in a real-world, community-based study in Bangladesh between January to April 2021. We compared both tests to the gold standard RT-PCR performed on a nasopharyngeal swab. We found the BD Veritor test to be more sensitive (78%) than the Standard Q (68%) in our study population.

The sensitivity of both rapid antigen tests was highly dependent on the Ct value of the specimen evaluated. Both tests were 100% sensitive among individuals with Ct < 20 – those with high viral loads. These findings are consistent with a study in China that reported 68% sensitivity and 100% specificity when a Ct value ≤40 was used as a cut off, as compared with 98% sensitivity and 100% specificity when Ct value≤ 30 (14).

Interestingly, we found that the sensitivity of both the rapid antigen tests was also high among asymptomatic individuals.. According to the RT-PCR analysis, the national prevalence rate of COVID-19 in Bangladesh during 26^th^ January–15^th^ March ranged from 2 to 8% after which the rate increased to 24% and peaked by beginning of April 2021. Of note, most of the asymptomatic RT-PCR positive participants were enrolled during the period in which infections surged, while most of the symptomatic individuals were enrolled prior to the surge. Therefore the performance of rapid antigen might vary among population groups due to different epidemiological, geographical conditions and impact of the variants of concern (VOC) circulating in that time period although no difference has been found until now (5)(15). In Bangladesh, it was the UK variant, followed by the he South Africa variant predominating in Bangladesh at the time (https://www.gisaid.org/).

The use of anterior nares specimens for the BD Veritor rapid antigen test was an added advantage, and study participants found this test to be more acceptable than the nasopharyngeal swab. An additional advantage of the BD Veritor testing was that it was carried out directly at the study site and results were available immediately. In several instances, study participants were informed to isolate themselves until RT-PCR results confirmed the infection. In addition, the analyzer provided with the kit was able to detect very faint bands on the device barely visible by the naked eye which reduces the chance of human bias. Inclusion of the positive and negative controls with other kit components was helpful for quality assessment of the analyzer before use.

A limitation of our study is that we compared rapid antigen tests that required different types of clinical specimens: For the BD Veritor we used anterior nares specimens for rapid test and a NP sample for PCR; the Standard Q uses only one nasopharyngeal specimen, which is diluted in viral transport media and used for both PCR and rapid test. If the concentration of the virus differs between AN and NP swabs, that is in the two sites of the nostril, it can also affect results. However, we did not see significant discrepancies between the two tests, except when specimens had high RT-PCR Ct values, indicating low viral loads. In that case both BD Veritor and the Standard Q failed to detect SARS-CoV-2 virus. Notably, we also found the BD Veritor rapid antigen test to be more sensitive even though the anterior nares specimen for it were collected later for the RT-PCR positive individuals.

Our study on rapid antigen tests is timely for Bangladesh which is currently experiencing a second wave and high rates of COVID-19 (16). Point of care tests are urgently needed for health facilities, travelers, workplaces and the general population, and our findings can help guide the implementation of these tests in Bangladesh. The BD Veritor test is sensitive enough to detect cases with high viral load in pre-symptomatic and early symptomatic cases as well as asymptomatic persons – groups that likely contribute to a significant proportion of transmission and spread of the disease (17). The patients who test positive by rapid antigen tests can readily be diagnosed with a minimum turnaround time, which offers the opportunity for early interruption of transmission through targeted isolation and contact tracing, as infectivity may be high with a Ct< 24 or so (18). Persons with negative rapid test who are suspected of having COVID can be tested by RT-PCR, depending on the epidemiologic context. Our information can provide implementation guidance when deciding testing strategies in different settings.

Many countries are now planning the expanded use of rapid antigen tests, and our results will provide guidance on their implementation in real-world settings such as that performed our study site in Bangladesh. Rapid antigen tests will be a critical component of COVID-19 control for the foreseeable future.

## Data Availability

All the data related to this manuscript are confidential. Hence, we are not able to make it public until our manuscript gets published in a peer reviewed journal.

## Acknowledgment

We gratefully acknowledge the support of Mass General Brigham (MGB) Center for COVID Innovation for providing the BD Veritor plus analyzer antigen detection kits and the Massachusetts General Hospital (MGH) for facilitation of the work. We thank the Bill and Melinda Gates Foundation for support to icddr,b investigators. icddr,b is grateful to the Governments of Bangladesh, Canada, Sweden, and the United Kingdom for providing core/unrestricted support. The overall support of Fondation Merieux to ideSHi is acknowledeged. We would like to express our gratitude to the participants for taking part and the field site staff of ideSHi for their effort to accomplish the work.

